# Migrant men and HIV care engagement in Johannesburg, South Africa

**DOI:** 10.1101/2023.08.23.23294266

**Authors:** Maria F. Nardell, Caroline Govathson, Sithabile Mngadi-Ncube, Nkosinathi Ngcobo, Daniel Letswalo, Mark Lurie, Jacqui Miot, Lawrence Long, Ingrid T. Katz, Sophie Pascoe

## Abstract

**Background:** South Africa (SA) has one of the highest rates of migration on the continent, largely comprised of men seeking labor opportunities in urban centers. Migrant men are at risk for challenges engaging in HIV care. However, rates of HIV and patterns of healthcare engagement among migrant men in urban Johannesburg are poorly understood.

**Methods:** We analyzed data from 150 adult men (≥18 years) recruited in 10/2020-11/2020 at one of five sites in Johannesburg, Gauteng Province, SA where migrants typically gather for work, shelter, transit, or leisure: a factory, building materials store, homeless shelter, taxi rank, and public park. Participants were surveyed to assess migration factors (e.g., birth location, residency status), self-reported HIV status, and use and knowledge of HIV and general health services. Proportions were calculated with descriptive statistics. Associations between migration factors and health outcomes were examined with Fisher exact tests and logistic regression models. Internal migrants, who travel within the country, were defined as South African men born outside Gauteng Province. International migrants were defined as men born outside SA.

**Results:** Two fifths (60/150, 40%) of participants were internal migrants and one fifth (33/150, 22%) were international migrants. More internal migrants reported living with HIV than men born in Gauteng (20% vs 6%, p=0.042), though in a multi-variate analysis controlling for age, being an internal migrant was not a significant predictor of self-reported HIV positive status. Over 90% all participants had undergone an HIV test in their lifetime. Less than 20% of all participants had heard of pre-exposure prophylaxis (PrEP), with only 12% international migrants having familiarity with PrEP. Over twice as many individuals without permanent residency or citizenship reported “never visiting a health facility,” as compared to citizens/permanent residents (28.6% vs. 10.6%, p=0.076).

**Conclusions:** Our study revealed a high proportion of migrants within our community-based sample of men and demonstrated a need for bringing PrEP awareness and services to migrants in Johannesburg. Future research is warranted to further disaggregate this heterogenous population by different dimensions of mobility and to understand how to design HIV programs in ways that will address migrants’ challenges.

## Introduction

South Africa has one of the highest rates of migration on the continent.^1^ In urban Gauteng Province alone, an estimated 29% of the population is “internal” migrants who have changed residences within the country and 6% is international migrants who have crossed country borders.^2^ Migration and mobility, both within and into South Africa, have been key drivers of the country’s HIV epidemic.^3,4^ While there are growing numbers of migrant women,^5^ the majority of migrants are men seeking work in urban centers.^6,7^ These men are often driven by a lack of employment opportunities^8^ and shifts in climate affecting agricultural practices in rural areas,^9–11^ and they often retain strong ties to their homes of origin.^8^ Migrant men in South Africa have borne twice the burden of HIV compared to non-migrant men with an HIV prevalence of ∼25.9%.^12^ Migrant men have been shown to be less likely to engage in HIV services, including HIV testing,^13–16^ pre-exposure prophylaxis (PrEP),^17,18^ and antiretroviral therapy (ART).^19,20^

The associations between migration and HIV acquisition, care engagement, and HIV-related outcomes are complex and incompletely understood.^5,21^ Some research has shown that people already living with HIV are more likely to migrate.^22,23^ Other evidence shows that some migrants may be healthier when they leave their homes of origin, often called the “healthy migrant” hypothesis.^24^ Migrants often have worse mental and physical health outcomes in their new destinations.^21,25^ Individuals who travel more frequently away from home have been shown to be at higher risk for acquiring HIV than those who travel less.^26^ In addition, migration and mobility pose significant challenges to health systems which assume a stable catchment area. South Africa’s decentralized health system is ill-suited for facilitating people changing clinics because it is difficult to easily transfer patient data across facilities.^27^ Migrants also face other intersecting psychosocial and structural challenges which affect HIV and healthcare engagement, such as the disruption of relocation,^22^ lifestyle changes,^5^ unstable or lack of employment,^21^ isolation and difficult living conditions,^25^ lack of residency or citizenship documentation,^28^ and fear and language issues.^29^ For male migrants, these challenges are compounded by internal and social constructs of masculinity, which impede care engagement in healthcare settings felt to be oriented towards women’s needs.^30^

The heterogenous nature of population mobility and varying definitions of migration further complicate efforts to understand migration and healthcare engagement, especially across different settings and populations.^31,32^ For example, some studies focus on the temporal nature of mobility (e.g., seasonal migration^33^), while others focus on specific social reasons for mobility (e.g., market traders^34^ or miners^35^), or spatial aspects of mobility, such as how distance traveled affects HIV acquisition risk.^36,37^ In addition, there are challenges accessing migrant men in research. For example, large trials to improve HIV testing uptake among men in South Africa have found substantial barriers to reaching men in communities with high mobility patterns.^38^ A demographic surveillance study from rural South Africa showed that 69% of the adult population cohort migrated out of the study area at least once during a 13 year follow-up period.^22^ Most of these men seek employment opportunities in cities in Gauteng Province, where Johannesburg is located, and yet research describing migrants within these urban destination sites remains limited and often comes from ethnographic rather than HIV studies.^39,40^

Movement into and within South Africa, particularly from rural to urban centers, continues to rise along with the country’s rapid socio-economic growth^22^ and accelerating urbanization throughout the region.^41^ There is a growing recognition of the need to make HIV care more accessible to migrants in order to achieve UNAIDS 95-95-95 goals.^30^ Yet despite migrants’ vulnerability to HIV acquisition^37,42^ and susceptibility to poor health engagement,^20,43,44^ there are few recent studies that seek to describe urban South African migrants, document their rates of HIV, or understand their health seeking behaviors.^45,46^ In this study, we sought to understand these gaps by characterizing migrant men within a broader sample of men recruited at community venues in Johannesburg for a study to understand men’s patterns of care engagement. We aimed to analyze participants’ burden of HIV, rates of HIV testing, awareness and knowledge of PrEP, and overall healthcare utilization. We hypothesized that both internal and international migrants, compared to men born in Gauteng, would be at a greater risk for acquiring HIV, and be less likely to engage in treatment and prevention services. Lastly, we hypothesized that migrants would be less likely than men born in Gauteng to have ever visited a health facility, and that migrants without citizenship or permanent residency would be less likely to have ever visited a health facility, as compared to citizens or permanent residents.

## Methods

This migration-specific study was embedded within our team’s broader “Men’s Choice” study, which included a prospective cohort of 150 adult men recruited at community-based, non-healthcare sites in Johannesburg in October and November 2020. The goal of the “Men’s Choice” study was to understand men’s preferences for accessing HIV and other healthcare services in Johannesburg through a discrete choice experiment (DCE) survey.^47^ The results of the DCE have been presented elsewhere.^47,48^ For this study, we administered a survey that preceded the DCE survey and included questions on men’s migration patterns, HIV care utilization, and socio-demographic measures.

## Participants, Study Setting, and Procedures

Five Johannesburg recruitment sites in the Hillbrow, Midrand, Woodmead and Roodeport areas were chosen following a period of observation and discussion with community stakeholders. These sites (factory, building materials store, taxi rank, homeless shelter, and public park) were selected to represent a range of locations where men go for work, transit, shelter, and leisure. We chose to focus on sites without pre-existing health programs in order to reach men who may not seek care at traditional healthcare settings. Recruitment occurred during the Covid-19 pandemic-related restrictions. Therefore, team members recruited participants by directly distributing study flyers at some sites (taxi rank, public park) or by asking leadership at some sites (homeless shelter, factory site, building materials store) to distribute flyers on the team’s behalf. These flyers gave information about the study and invited men to contact the study team if they were interested in participating. Those men who made contact were then given further information and taken through the consent process over the phone. The team checked telephone numbers, names and identifiers to ensure that the same participant did not enroll twice. Several of the first survey interviews were completed over the phone, but as restrictions lifted in later 2020, survey interviews were then completed face-to-face. After completing the survey, participants were reimbursed for their time with a ZAR150 (∼USD10) electronic shopping voucher, sent directly to the participant’s cell phone number.

## Eligibility

We included men who were 18 years or older, willing to participate in a 2-hour survey (including the DCE), and able to provide informed consent. Given Covid-19 pandemic restrictions, participants were also required to have access to a cell phone number so that informed consent and, if necessary, survey interviews could be conducted telephonically. Men were excluded from the study if they were believed to be intoxicated at the time of consent (in which case, they were invited to return the following day), unable to understand the study information, or had previously enrolled in the study.

## Measures and Study Design

We used the Theory of Triadic Influence to inform our selection of measures in the socio-demographic survey in order to understand correlates of HIV-related behavior, focusing on individual, interpersonal, and structural factors important for positive behavior change.^49^ In order to understand the role of migration and mobility in this population, questions in the socio-demographic survey included location of birth (South African province or country outside South Africa), the location where participants spend the majority of their time, duration of time in Gauteng, if and where participants moved in the past two years (between districts, provinces, or countries), reason for moving in the past two years, if applicable, and current residency status in South Africa. The list of socio-demographic survey measures included is shown in Appendix Table 1. Measures included the frequency of, location of, and reasons for health facility visits; ever testing for HIV and frequency of testing for HIV; self-reported HIV status; and awareness and knowledge of PrEP.

**Table 1.** Socio-demographic characteristics of participants by migrant status (Total N=150)

|  | All migrants |  | Internal migrants |  | International migrants |  | Men born in Gauteng Province |  |
| --- | --- | --- | --- | --- | --- | --- | --- | --- |
| Age (mean [SD]) | 32 [10.3] |  | 38 [9.5] |  | 39 [12.0] |  | 31 [10.3] |  |
|  | N | Column % | N | Column % | N | Column % | N | Column % |
| Participants | 93 | 62.0 | 60 | 40.0 | 33.0 | 22.0 | 57 | 0.38 |
| Province or country of origin |  |  |  |  |  |  |  |  |
|  |  |  | Limpopo 22 | 36.7 | Zimbabwe 25 | 75.8 |  |  |
|  |  |  | KwaZulu-Natal 10 | 16.7 | Mozambique 4 | 12.1 |  |  |
|  |  |  | Eastern Cape 8 | 13.3 | Ghana 1 | 3.0 |  |  |
|  |  |  | Free State 7 | 11.7 | Lesotho 1 | 3.0 |  |  |
|  |  |  | Mpumalanga 7 | 11.7 | No response 2 | 6.1 |  |  |
|  |  |  | North West 3 | 5.0 |  |  |  |  |
|  |  |  | Western Cape 2 | 3.3 |  |  |  |  |
|  |  |  | Northern Cape 1 | 1.7 |  |  |  |  |
| Educational level |  |  |  |  |  |  |  |  |
| Primary | 8 | 8.6 | 4 | 6.7 | 4 | 12.1 | 3 | 5.3 |
| Secondary | 70 | 75.3 | 50 | 83.3 | 20 | 60.1 | 48 | 84.2 |
| University | 6 | 6.5 | 4 | 6.7 | 2 | 6.1 | 4 | 7 |
| Vocational | 9 | 9.7 | 2 | 3.3 | 7 | 21.2 | 2 | 3.5 |
| Relationship status |  |  |  |  |  |  |  |  |
| Living with partner | 27 | 29.0 | 13 | 21.7 | 14 | 42.4 | 13 | 22.8 |
| Not living with partner | 36 | 3.9 | 24 | 40.0 | 12 | 36.4 | 24 | 42.1 |
| No relationship | 30 | 3.2 | 23 | 38.3 | 7 | 21.2 | 20 | 35.1 |
| Employment status |  |  |  |  |  |  |  |  |
| Student | 2 | 2.1 | 2 | 3.3 | 0 | 0 | 2 | 3.5 |
| Have an employer | 4 | 4.3 | 4 | 6.3 | 0 | 0 | 5 | 8.8 |
| Odd jobs/piece work | 8 | 8.6 | 2 | 3.3 | 6 | 18.2 | 2 | 3.5 |
| Self-employed | 13 | 14.0 | 3 | 5.0 | 10 | 30.3 | 10 | 17.5 |
| Unemployed | 66 | 71.0 | 49 | 81.7 | 17 | 51.5 | 37 | 64.9 |
| Retired | 0 | 0.0 | 0 | 0.0 | 0 | 0 | 1 | 1.8 |
| Duration of time living in Gauteng |  |  |  |  |  |  |  |  |
| <1 year | 14 | 15.1 | 11 | 18.3 | 3 | 9.1 | 3 | 5.3 |
| 1-2 years | 8 | 8.6 | 5 | 8.3 | 3 | 9.1 | 0 | 0 |
| 3-10 years | 29 | 31.2 | 15 | 25.0 | 14 | 42.4 | 4 | 7 |
| >10 years | 42 | 45.2 | 29 | 48.3 | 13 | 39.4 | 50 | 87.7 |
| Lived outside Gauteng in the past two years | 15 | 16.1 | 11 | 18.3 | 4 | 12.1 | 5 | 8.8 |
| Reason for coming to Gauteng in the past two years (total N=18) |  |  |  |  |  |  |  |  |
| Work opportunity | 8 | 44.4 | 5 | 45.5 | 3 | 75 | 2 | 67.7 |
| Education opportunity | 2 | 11.1 | 2 | 18.2 | 0 | 0 | 0 | 0 |
| Family reason | 5 | 27.8 | 4 | 36.4 | 1 | 25 | 1 | 33.3 |

## Statistical Methods

We defined internal migrant as South African men born outside of Gauteng Province, and international migrants as men born outside South Africa. We chose to define migrants based on their location of birth to broadly capture men who traveled to Gauteng Province at some point in their lifetime. The International Organization for Migration similarly defines the term “migrant” broadly to encompass “the common lay understanding of a person who moves away from his or her place of usual residence, whether within a country or across an international border, temporarily or permanently, and for a variety of reasons.”^50^ We distinguished between men born within and outside of South Africa because these groups may have different HIV and other healthcare experiences in South Africa based on factors such as xenophobia,^51^ language,^19^ identification documentation, and citizenship status.^28^ We also recognized that internal migrants and international migrants are both heterogenous groups. Therefore, we sought to measure other aspects of migration and mobility, including length of time in Johannesburg, reasons for moving, travel in the past two years, and residency status.

We estimated the prevalence of all migrants, internal migrants, and international migrants by calculating the proportion of each of these categories among all participants. We used Fisher’s exact tests to assess associations between migration factors and HIV and health utilization outcomes. Specifically, we compared self-reported HIV status, HIV testing in the past year, and PrEP knowledge between internal migrants and men born in Gauteng and between international migrants and men born in Gauteng. We also compared the proportion of men ever visiting a health facility between South African citizens or permanent residents and men without permanent residency or citizenship. In addition, we used logistic regression to predict HIV status by migration history, controlling for age.

### Ethics

The ethics committees at the University of the Witwatersrand (M191068), Mass General Brigham (Harvard University) (2020P002251), and Boston University (H-40529) approved the study. All participants provided written informed consent. Study data were collected and managed using Research Electronic Data Capture (REDCap), a secure, web-based tool.

## Results

### Socio-Demographic Characteristics of Migrants

In our sample of 150 men, nearly two thirds (62%) were migrants, as shown in Table 1. Two fifths of these migrants were internal migrants and one fifth were international migrants. The internal migrants represented all South African provinces outside of Gauteng, with the highest proportion born in Limpopo, followed by KwaZulu-Natal. All international migrants reported being from other countries on the African continent, with four fifths from Zimbabwe. Migrants ranged in age from 20 to 73 years, with a mean age of 39 years. The majority (85%) of all migrants reported living in Gauteng Province for more than one year, and just under half of all migrants had lived in Gauteng for more than ten years. Of the migrants who reported on their reason for moving to Gauteng, over half cited seeking or finding employment, followed by family reasons or an educational opportunity. Most migrants were in a relationship, but the majority did not live with their primary partner. The vast majority (93%) of international migrants did not have visas to live and work in South Africa.

### HIV Status

Out of the 137 participants who chose to report their HIV status, 11 internal migrants, 3 international migrants, and 3 non-migrants reported living with HIV, as shown in Figure 1. More internal migrants reported living with HIV (20.4%) versus men born in Gauteng (5.8%) (p=0.042). However, in a multi-variate analysis, being an internal migrant was not a significant predictor of self-reported HIV positive status, controlling for age. There was no difference in self-reported HIV status between international migrants and men born in Gauteng.

**Figure 1.**
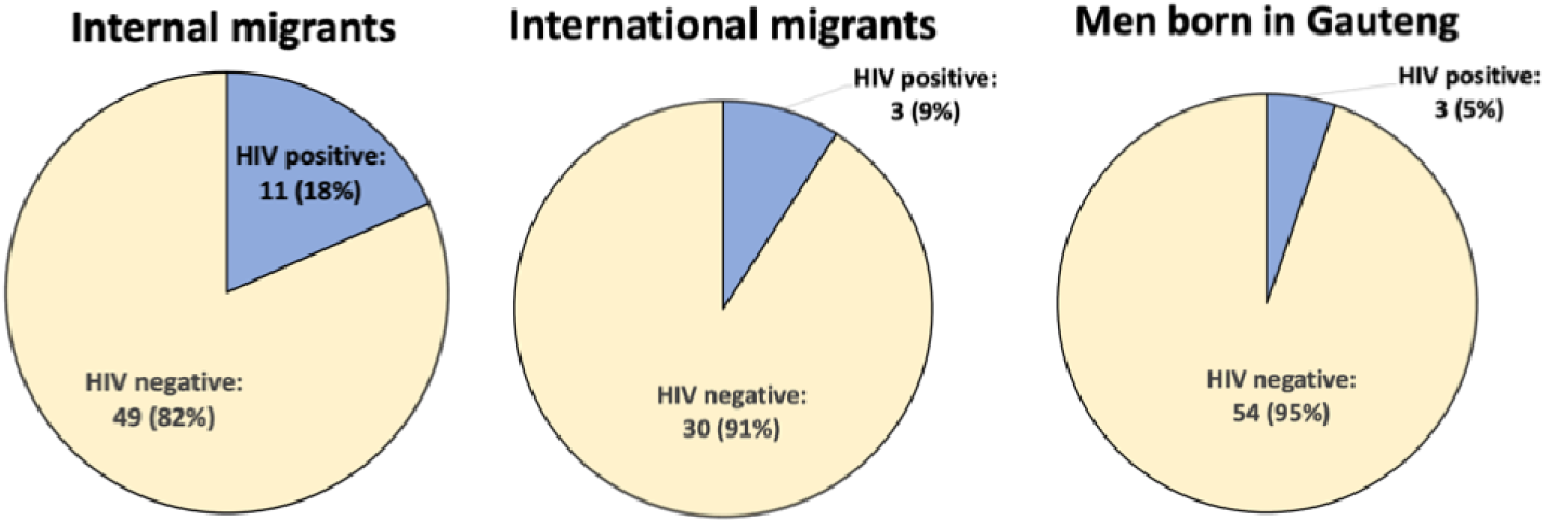
Self-reported HIV status among participants. Using a Fisher’s exact test to compare HIV status between internal migrants and men born in Gauteng Province, more internal migrants reported living with HIV 11/54 (20.4%) versus non-migrant men 3/52 (5.8%)

### HIV Care Cascade Engagement and PrEP Knowledge

Almost all participants reported ever testing for HIV (94.6%), with no statistically significant difference in ever testing between men born in Gauteng as compared to either migrant group. (See Table 2) Nearly two thirds of all men tested in the past year, again with no difference between men born in Gauteng as compared to either migrant group. Most men reported “No” or “Unsure” when asked if they had heard of PrEP (84.7%), with similar proportions of internal migrants and international migrants reporting this. When asked what PrEP does, nearly a third of non-migrants answered correctly that it prevented HIV, versus 20.4% internal migrants and 12% of international migrants who answered correctly.

**Table 2.**
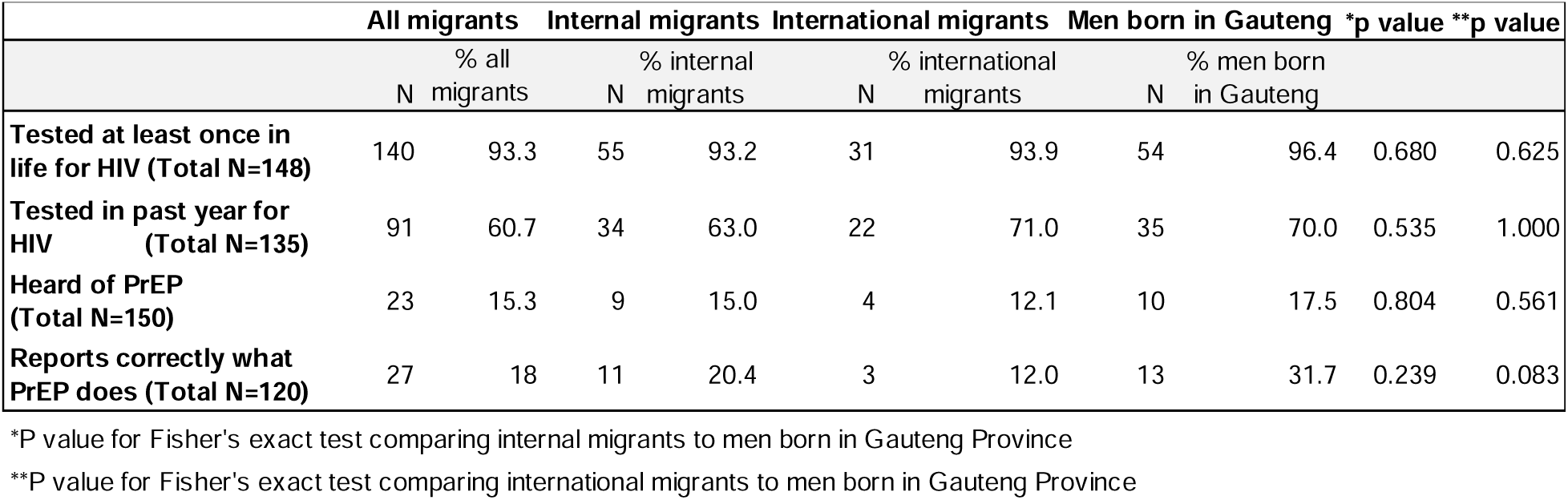
HIV testing and PrEP knowledge, comparing migrants to men born in Gauteng Province.

### Residency Status and Overall Healthcare Utilization

As shown in Table 3, among the international migrants who reported on their residency status, the majority were not permanent residents or citizens. Among all men who reported on their residency status as well as their healthcare utilization, 28.6% men without permanent residency reported never visiting a health facility as compared to 10.6% citizens/permanent residents (p=0.076). The proportions of migrants and men born in Gauteng who reported that they had ever visited a health facility were similar.

**Table 3.** Healthcare utilization among all participants by residency status and migrant status. We used Fisher’s exact tests to compare ever having visited a health facility by residency status and by migrant status, finding higher proportions of men without citizenship or permanent residency who reported never having visited a health facility.

|  | Permanent residents or citizens |  | Migrants without citizenship or permanent residency |  |  |
| --- | --- | --- | --- | --- | --- |
|  | N | % all permanent residents/citizens | N | % men without citizenship or permanent residency | P-value |
| **Residency status (N=144\*)** | 119** | N/A | 25 | N/A | N/A |
| **Ever visited a health facility (Total N=106)** | 76 | 89.4 | 15 | 71.4 | 0.076 |
|  | **All migrants** |  | **Men born in Gauteng Province** |  |  |
|  | N | % all migrants | N | % men born in Gauteng Province | P-value |
| **Ever visited a health facility (Total N=112)** | 58 | 84.1 | 37 | 86.0 | 1.000 |

\*27 out of 33 international migrants reported on their residency status in South Africa.
\*\*Two international migrants reported that they had a permanent residency status in South Africa.

## Discussion

In this study of men in Johannesburg, we found that migrants comprised approximately two thirds of our sample, with nearly twice as many internal migrants as compared to international migrants. Almost three times as many internal migrants reported they were living with HIV as compared to men from Gauteng, and older men were both more likely to be internal migrants and more likely to be living with HIV. Rates of ever testing for HIV were high for all participants, though all participants had low awareness and knowledge of PrEP. Most men reported visiting a health facility, although almost a third of migrants without South African permanent residency or citizenship reported never visiting a health facility at any point in their lifetime.

While our study was not designed to representatively sample migrants within Johannesburg, our finding of a high proportion of internal migrants from Limpopo may support evidence that economic opportunity remains a major driver of migration and mobility within South Africa.^36,52,53^ Limpopo is South Africa’s fifth most populous province,^54^ but it has one of the highest rates of poverty in the country.^55^ Our high representation of international migrants from Zimbabwe is unsurprising given that there are an estimated one million Zimbabweans in South Africa who migrate for factors including economic opportunities and political unrest in their home country.^53^ Yet over 80% internal migrants and over half of the international migrants in our sample reported having no current formal or informal (e.g., temporary “piece work” arrangements with payment by unit produced) employment. Despite Johannesburg’s reputation as the “City of Gold,” the economic landscape for migrants remains challenging, with problems including persistent electricity cuts and job losses in the construction, manufacturing, and mining industries.^56,57^ International migrants may face additional hurdles due to new South African laws restricting employment of foreign workers.^58^ In addition, Covid-related restrictions during the time of this study further tightened job opportunities and exacerbated vulnerabilities for those working in South Africa’s informal labor sector.^59,60^

We found that older men were both more likely to be internal migrants and more likely to be living with HIV. Prior research has supported the association between migration and HIV risk and prevalence,^5,26,37^ as well as between older men and HIV infection.^61–63^ Data also show that older men may be less likely to test^64–66^ and less likely to know their HIV status as compared to younger men.^67^ Given that we relied upon self-report for determining HIV status in this study, our data may have under-estimated the proportion of participants living with HIV, particularly for older migrant men. Our finding of high rates of ever testing for HIV among all participants may reflect South Africa’s overall gains in HIV testing.^68^ Yet this national progress has been uneven among different populations.^69^ Approximately a third of participants had not tested for HIV in the past year, despite the fact that testing at least annually has been shown to reduce the burden of undiagnosed HIV,^13^ and testing remains an important entry point for engagement in HIV care.^70–72^ However, it may be difficult to interpret these data in the setting of Covid-related restrictions, which likely had an impact on HIV testing and other healthcare engagement for all participants during the year preceding this study.^59^

Despite overall high rates of ever HIV testing, all participants in our cohort had low awareness of and low knowledge of PrEP. This may be suggestive of general gaps in PrEP awareness^73,74^ and knowledge^75^ in South Africa, despite South Africa’s efforts to promote PrEP use through existing services and differentiated service delivery models.^76^ In addition, although migrant men are considered a key population in need of targeted HIV services,^77,78^ there are few PrEP programs reaching men more broadly,^79,80^ and none to our knowledge targeting migrant men specifically. In addition, more than a quarter of men without permanent residency or citizenship reported never visiting a health facility. While it is South African policy that healthcare should be provided in the public sector to all who need it, barriers to healthcare for non-nationals may include fear of exposing one’s undocumented status, stigma for being a non-citizen, or the potential for healthcare fees.^28^

Taken together, our findings have important implications for the design of health and HIV programs in South Africa, as well as for other countries with growing rates of migration. In order to close the remaining gaps in reaching UNAIDS targets,^81^ it is imperative to engage migrant men as a key population. In particular, there is a need to better understand and address migrants’ knowledge of and attitudes toward PrEP^45,46^ in order to design PrEP programs that are more inclusive of them.^77,82^ Existing research on HIV care for migrants in Johannesburg has focused on international migrants,^28,83^ who receive justified national and international attention for challenges that may include xenophobia,^84^ language barriers, and lack of permanent residency or citizenship status.^85,86^ However, our study also highlights the vulnerabilities and barriers to care that may be faced by internal migrants (e.g., difficulties transferring between clinics without transfer documentation^87^), who may warrant targeted attention to ensure that they are reached by HIV services.

Our study is cross-sectional and thus limited in that it represents a population of migrant men at one point in time. In addition, our study recruited from community locations in Johannesburg to find men who may not commonly access healthcare, so our findings may not be generalizable to migrants in other parts of the province or country. We defined “migrant” as born in a province or country outside of Gauteng. This approach, focusing on the spatial dimension of migration, is similar to what has been used in several other studies involving migrants.^12,19,88^ However, we acknowledge inherent limitations to this approach, as temporal dimensions of mobility (e.g., the amount of time spent in a particular location or seasonal travel) and social dimensions of mobility (e.g., the reason for travel) may contribute to a more nuanced understanding of migrant and mobile populations.^7,32,37^ We conducted our study in English, isiZulu, Setswana, and isiXhosa, and Sesotho, but we were unable to use all potential first languages for migrants. Response rates were variable for some survey items, which limits our ability to interpret these data. In particular, data on ART use were not obtained due to a data collection error, so we were unable to assess this aspect of care engagement for participants living with HIV. This study focused on HIV and overall healthcare engagement, but future research may consider more deeply the impact of trauma and mental health care as an important need within this population.^89^

## Conclusions

Our study revealed a high proportion of migrants within our community-based sample of men and highlighted a need for bringing PrEP services to migrant men in Johannesburg. Future research is warranted to further disaggregate this heterogenous population by different dimensions of mobility and to understand how to design HIV prevention programs in ways that will address migrants’ challenges. We anticipate that our findings, as well as further research among different migrant populations, will have relevance not only for South African’s health system and policies, but also for other African countries experiencing quickly growing rates of migration and urbanization.^90^

## Appendices

**Appendix Table 1:**
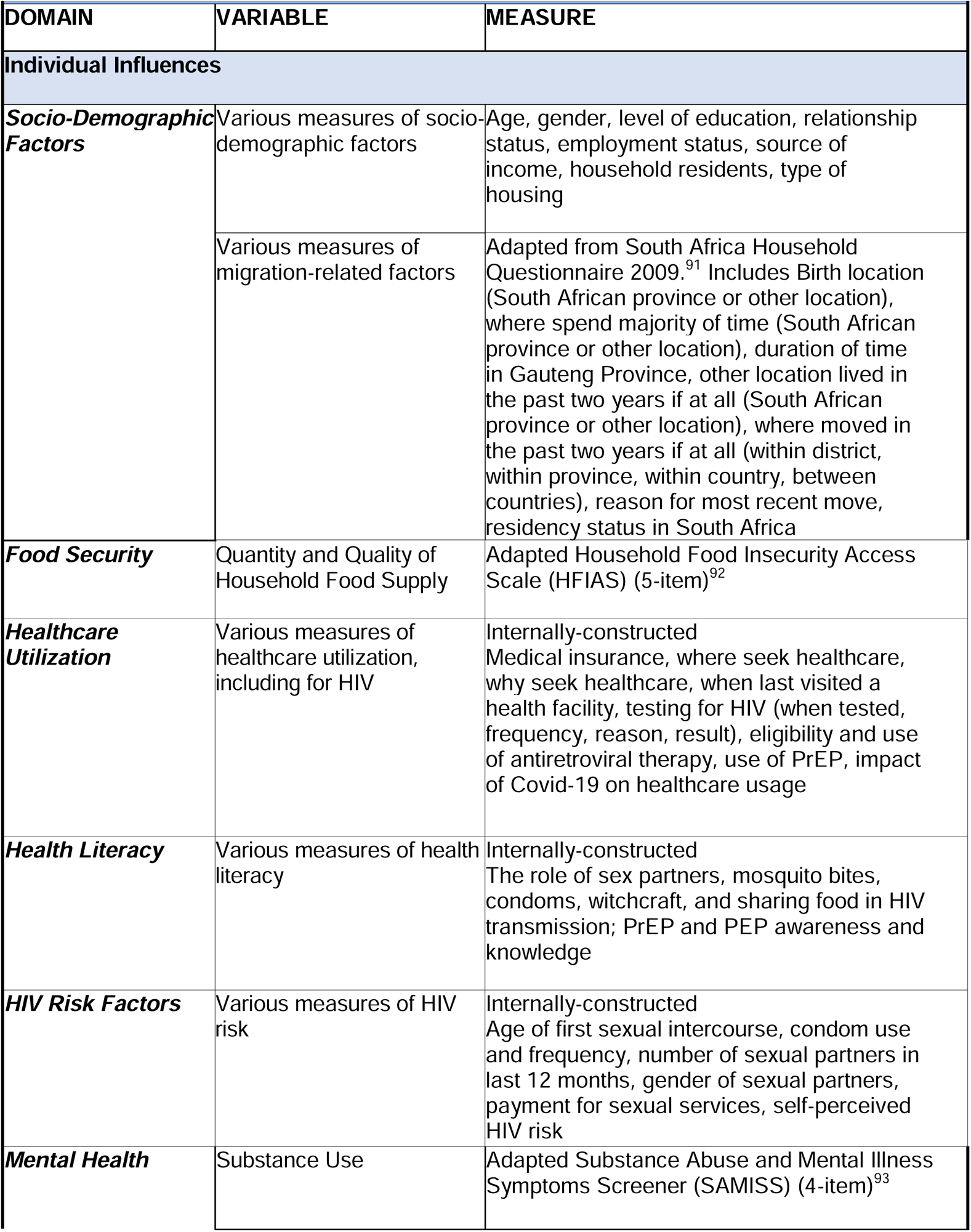
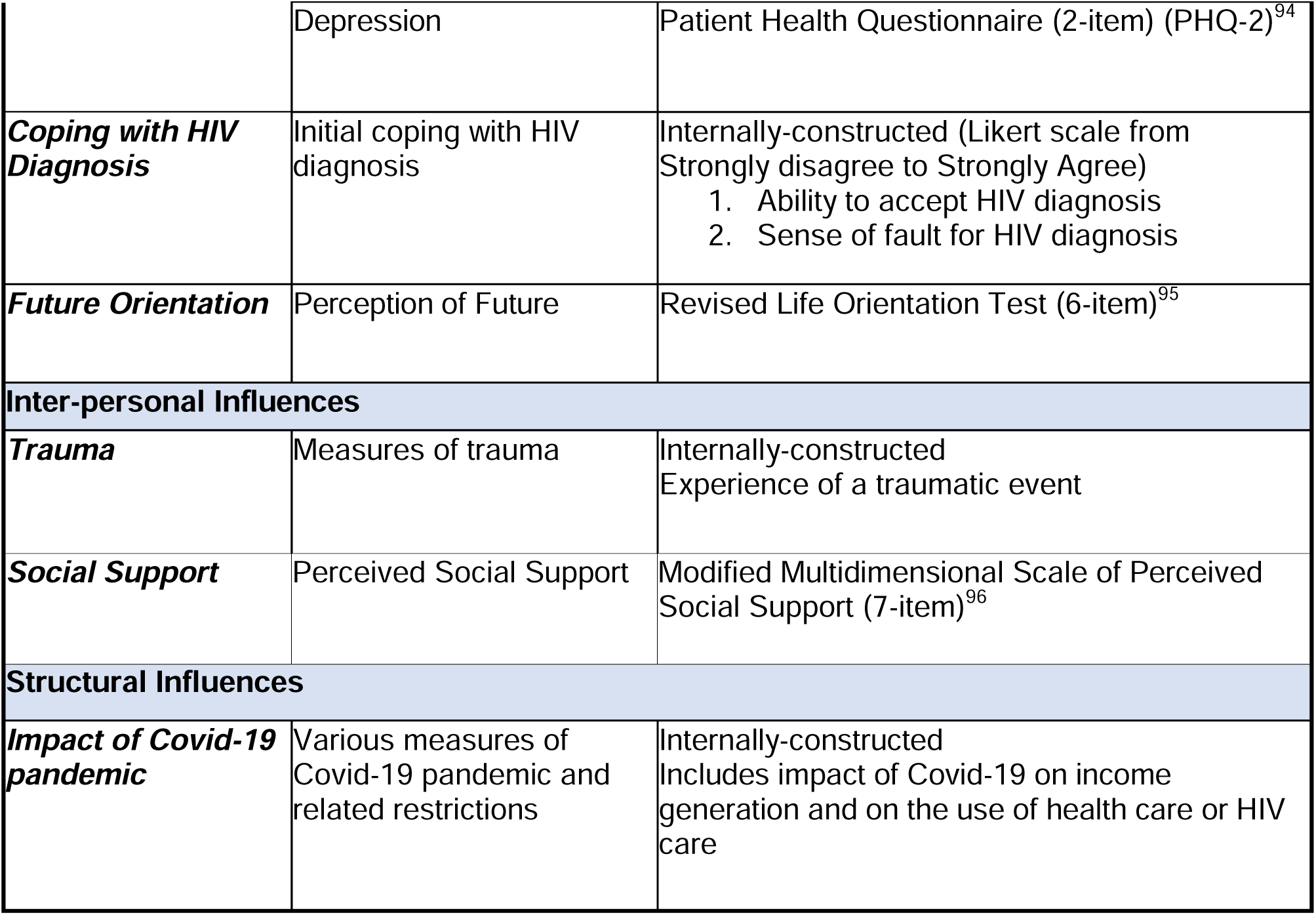
Key Domains, Variables, and Measures.

## Data Availability

All data produced in the present study are available upon reasonable request to the authors.

